# Consideration of within-patient diversity highlights transmission pathways and antimicrobial resistance gene variability in vancomycin resistant *Enterococcus faecium*

**DOI:** 10.1101/2022.09.23.22279632

**Authors:** Martin P McHugh, Kerry A Pettigrew, Surabhi Taori, Thomas J Evans, Alistair Leanord, Stephen H Gillespie, Kate E Templeton, Matthew TG Holden

## Abstract

**Background:** Whole genome sequencing (WGS) is increasingly applied to healthcare-associated vancomycin-resistant *Enterococcus faecium* (VREfm) outbreaks. Within-patient diversity could complicate transmission resolution if single colonies are sequenced from identified cases.

**Objectives:** Determine the impact of within-patient diversity on transmission resolution of VREfm

**Methods:** Fourteen colonies were collected from VREfm positive rectal screens, single colonies were collected from clinical samples, and Illumina WGS performed. Two isolates were selected for Oxford Nanopore sequencing and hybrid genome assembly to generate lineage-specific reference genomes. Mapping to closely related references was used to identify genetic variations and closely related genomes. A transmission network was inferred for the entire genome set using Phyloscanner.

**Results:** In total, 229 isolates from 11 patients were sequenced. Carriage of 2-3 sequence types was detected in 27% of patients. Presence of antimicrobial resistance genes and plasmids was variable within genomes from the same patient and sequence type. We identified two dominant sequence types (ST80 and ST1424), with two putative transmission clusters of two patients within ST80, and a single cluster of six patients within ST1424. We found transmission resolution was impaired using fewer than 14 colonies.

**Conclusions:** Patients can carry multiple sequence types of VREfm, and even within related lineages the presence of mobile genetic elements and antimicrobial resistance genes can vary. VREfm within-patient diversity should be considered to ensure accurate resolution of transmission networks.

## INTRODUCTION

*Enterococcus faecium* is a leading nosocomial pathogen causing opportunistic infections mostly in immunocompromised hosts. Antimicrobial resistance is a key concern, particularly against front-line anti Gram-positive agents amoxicillin and vancomycin.^1^ Vancomycin-resistant *E. faecium* (VREfm) infections lead to increased length of stay, cost an estimated USD200 per case per day, and confer mortality of 23-47%.^2–7^ In 2020, vancomycin resistance of 45.6% was reported among all *E. faecium* bloodstream isolates in Scotland, among the highest rates in Europe.^8^

In healthcare institutions asymptomatic intestinal carriage of VREfm can lead to shedding into the environment and transfer to other patients or staff, challenging efforts to limit the incidence of nosocomial infections.^9^ Whole genome sequencing (WGS) is increasingly applied to investigate transmission networks and identify control measures.^10,11^ Many WGS based analyses of bacterial outbreaks, however, rely on analysing single colony picks from clinical samples assuming that this represents the entire infecting or colonizing population within individual patients.^12^ It is increasingly recognised that within-patient diversity of bacterial populations can be significant and can influence transmission network resolution.^13–19^ Several studies have identified that individual patients can carry multiple strains of *E. faecium* concurrently, but few have applied this to transmission resolution.^20–24^

In this study, we aimed to identify within-patient diversity of VREfm from rectal screening swabs and determine how this impacts transmission inference in a 1-month snapshot on a haematology unit. We designed a sampling strategy to reliably detect within-patient diversity and supplemented short-read and long-read sequencing to generate high-quality reference genomes to identify genomic variants in the isolate collection.

## MATERIALS AND METHODS

### Isolates

Rectal swabs were collected at admission and on all inpatients on the haematology unit developing febrile neutropenia (neutrophils <0.9×10^9^/l or <1.0×10^9^/l and falling after chemotherapy, plus body temperature ≥38°C). Swabs were plated to CHROMID® VRE agar (bioMérieux, Marcy-l’Étoile, France), species identification and vancomycin resistance were confirmed with MALDI-TOF (Microflex instrument, Bruker, Billerica, USA) and VITEK-2 (bioMérieux) with EUCAST breakpoints. All purple colonies from VREfm positive plates were stored at −80°C in a Microbank cryovial (Pro-Lab Diagnostics, Birkenhead, UK). Any VREfm isolated from clinical samples within 60 days of a rectal positive were also stored. Patient metadata was retrieved from electronic records and movements visualised with HAIviz v0.3 (https://haiviz.beatsonlab.com/). This work was approved by the NHS Scotland BioRepository Network (Ref TR000126) and the University of St Andrews Research Ethics Committee (Ref MD12651).

### Genome Sequencing

Cryovials were re-plated on CHROMID® VRE agar, and fourteen random purple colonies were incubated overnight in 5 ml brain heart infusion broth (Oxoid). Cells were pelleted and DNA extracted using the DNA Mini kit on a QiaSymphony instrument (Qiagen).

Short read libraries were prepared using the Nextera XT kit (Illumina, San Diego, USA) and sequenced with a MiSeq instrument (Illumina) using 300 bp paired-end reads on a 600-cycle v3 reagent kit.

For long read sequencing, isolates VRED06-02 (ST1424) and VRED06-10 (ST80) were selected at random from the first sample with multiple sequence types detected (sample VRED06 from patient P49). Long read libraries were generated with the LSK109 Ligation Sequencing Kit (Oxford Nanopore Technologies, Oxford, UK) and sequenced for 8h using an R9.4 flow cell on a GridION sequencer (Oxford Nanopore Technologies) with high accuracy basecalling in MinKNOW v19.12.6 (Oxford Nanopore Technologies).

Sequence data from this study have been deposited in the NCBI under BioProject accession number PRJNA877253 (https://www.ncbi.nlm.nih.gov/bioproject/PRJNA877253).

### Sequence Assembly and Mapping

Short reads were quality trimmed with Trimmomatic v0.32.^25^ MLSTs were determined with SRST2 v0.2.0^26^ and the *E. faecium* pubMLST database.^27^ A core alignment was generated by mapping short reads to the reference genome(s) with Snippy v4.6.0 default settings (https://github.com/tseemann/snippy) and masking all putative transposases, prophage regions, and recombination blocks. Recombination blocks were identified with Gubbins v2.4.1.^28^ Non-ACGT bases were converted to N with snippy-clean and a core SNP alignment generated with snp-sites v2.5.1.^29^ The 130 ST80 genomes mapped to VRED06-10 generated an initial alignment of 2,814,943 bases, 202,738 bases were masked, and the final alignment contained 96 variant sites; the 97 ST1424 genomes mapped to VRED06-02 generated an initial alignment of 2,945,113 bases, 227,540 bases were masked, and 13 variant sites remained. Maximum-likelihood phylogenies were constructed with IQTree v2.0.3 with automatic model selection and 1000 ultrafast bootstraps.^30–32^ Phylogenies were visualised with iTOL.^33^ Short read assemblies were generated with Unicycler v0.4.8^34^ and searched for antimicrobial resistance genes using Abricate v1.0.1 (https://github.com/tseemann/abricate) with default settings and the ResFinder database.^35^

Long reads <1000 bp were removed with Nanofilt v2.7.1^36^ and adapters trimmed and chimaeras split with Porechop v0.3.2 (https://github.com/rrwick/Porechop). Reads were split into 12 subsamples and three assemblies made with four different assemblers: Flye v2.8.1, Redbean v2.5, Raven v1.1.10, and Miniasm v0.1.3,^37–40^ giving 12 assemblies in total. A consensus assembly was generated with Trycycler v0.3.3^41^ and polished with Medaka v0.11.5 (https://github.com/nanoporetech/medaka) and 2-3 cycles of Pilon v1.23.^42^ Assembly quality was assessed with assembly-stats v1.0.1 (https://github.com/sanger-pathogens/assembly-stats), Ideel (https://github.com/phiweger/ideel), and Busco v4.1.4.^43^

Polished assemblies were annotated with Prokka v1.14.6^44^ using the Aus0004 reference genome (Accession CP003351) with the --proteins option. Abricate identified matches to ResFinder, VirulenceFinder, and PlasmidFinder databases^45–48^ and putative prophages were identified with PHASTER.^49^ Plasmid copy numbers were estimated using short reads and Snippy: average depth for each plasmid was divided by the average depth of the chromosome.

Plasmids in the polished assemblies were compared to each other with Mash v2.2.2.^50^ To detect plasmids, those present in the two polished assemblies were used as references against all short read sets in Snippy and considered present if ≥85% bases were called with <20 SNPs/1000 bp.^51^

### Transmission Network Inference

All short reads were mapped to the VRED06-10 ST80 reference chromosome with Snippy, the V24 *E. faecium* ST80 genome (Accession CP036151) was included as an outgroup. An alignment of 2,814,943 bases was generated and 1,418,409 bases masked as above. A posterior set of phylogenies were generated with MrBayes v3.2.7.^52^ Two MCMC runs of four coupled chains were run for 5,000,000 generations, sampling every 5000. The final standard deviation of split frequencies was 0.013, the log-likelihood was stable, and the effective sample size of all parameters was >800. A random sample of 100 posterior trees was input to Phyloscanner v1.6.6.^53^ Sankoff parsimony reconstruction was performed with *k* parameter of 281494.5, equivalent to a within-patient diversity threshold of 10 SNPs as used in other studies.^54^ A transmission network was constructed in Cytoscape v3.9.0^55^ showing edges with complex or transmission state and >0.5 probability. The role of smaller numbers of colony picks on transmission resolution was investigated by repeating the above with the first 3, 5, and 10 genomes per sample.

### Statistical Analysis

To determine the optimal number of colonies to analyse for within-sample diversity a power calculation was performed as described by Huebner et al:^56^

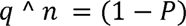

Where *q* = 1 – concentration of organisms, ^ = exponentiation operator, *n* = number of colonies sequenced, and *P* = probability of finding one or more variants.

Population variants were considered distinct if they differed by >10 chromosomal SNPs.^54^ Moradigaravand *et al*^20^ show rectal VREfm populations harbour minority variants at 20-50% of the total population based on sequencing ∼5 colonies. However, no minor population variants were identified in blood cultures based on sequencing ∼10 colonies.^20^ For rectal samples, we determined 14 colonies per sample would detect a variant at 20% of the population with 95% confidence, and for blood cultures we sequenced single colonies to identify the sequence type causing invasive disease.

Presence/absence matrices of antimicrobial resistance (AMR) genes were generated in R v4.0.5 using ggplot2 and patchwork packages.^57–59^

## RESULTS AND DISCUSSION

### Epidemiological Context

This study was performed over one month in 2017 on an inpatient unit for haematological malignancies, split into two wards (A and B). VREfm rectal screening was performed on all new admissions and any inpatients with febrile episodes to inform patient placement and antimicrobial administration. There was significant overlap between patient stays with some patients moving between the two study wards or to other wards in the hospital (Figure 1). Patients were cohorted or placed in single rooms when colonised with VREfm or other alert pathogens. However, not all rooms had ensuite bathroom facilities so risk of VREfm transmission remained. At the time of the study, surveillance systems in the hospital had not detected any suspected VREfm outbreak within the study population.

**Figure 1.**
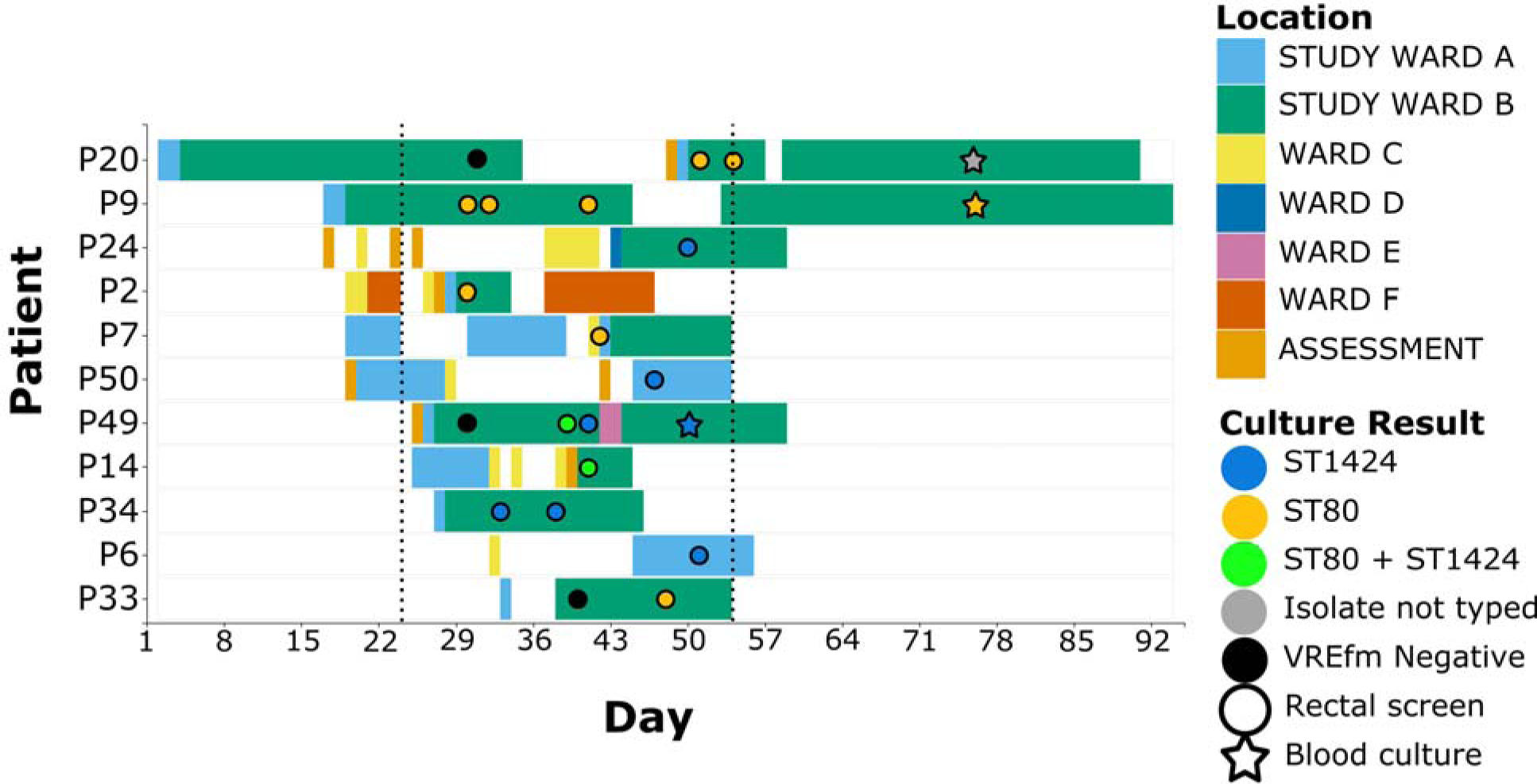
Patient timeline. Each row denotes the location of a patient during admission, blocks denote hospital stay, circles denote VREfm cultures, stars denote bloodstream isolates, dotted lines indicate the start and end of prospective collection of screening isolates for this study. This study was undertaken mainly within Wards A and B, although patients were moved to different wards within the hospital during their stay and were often admitted through the assessment unit.

### Results of VREfm Screening

In total, 45 rectal swabs from 27 patients were screened for VREfm. Of these, 18 samples from 13 patients were VREfm positive (Table 1). Three (23.1%) colonised patients developed VREfm bacteraemia 9, 24, or 46 days after being identified as VREfm carriers. We applied our sampling strategy to 16 rectal screens generating 224 isolates from 11 patients, and five blood cultures (five isolates) from two patients yielding a total of 229 isolates. Two rectal swabs and one blood culture were not available for further study. Most patients were female, the median age was 66 years, and a range of primary diagnoses were present (Table 1). Most colonised patients had received antibiotics in the preceding six months and 30% had received vancomycin (Table 1).

**Table 1.** Characteristics of patients with rectal VREfm colonisation (n = 13)

| **Demographics** | **N (%)** |
| --- | --- |
| Female | 8 (61.5) |
| Age, median (range) years | 66 (37-77) |
| Primary diagnosis |  |
| Acute myeloid leukaemia | 3 (23.1) |
| Diffuse large B cell lymphoma | 3 (23.1) |
| Multiple myeloma | 3 (23.1) |
| Myelodysplasia | 2 (15.4) |
| Chronic lymphocytic leukaemia | 1 (7.7) |
| Mantle cell lymphoma | 1 (7.7) |
| Antimicrobial administration |  |
| Any antibiotics in the 7d prior to positive screen | 12 (92.3) |
| Any antibiotics in the 6m prior to positive screen | 12 (92.3) |
| Vancomycin in the 7d prior to positive screen | 1 (7.7) |
| Vancomycin in the 6m prior to positive screen<sup>a</sup> | 3 (30.0) |
| Outcomes within 60d of VREfm positive screen |  |
| VREfm bloodstream infection | 3 (23.1) |
| ICU admission | 1 (7.7) |
| Death | 0 (0) |

<sup>a</sup> Information available for 10 patients

### Simultaneous carriage of multiple VREfm strains

*In silico* MLST typing using short reads from all 229 genomes showed ST80 (n=130), ST1424 (n=97), ST789 (n=1), and ST1659 (n=1) from the hospital-associated clade A1^60^ were present (Table 2). Multiple STs were detected in three (27%) samples. Sample VRED06 from patient P49 contained 10 (71.4%) ST80, three (21.4%) ST1424, and one (7.1%) ST789 isolate; sample VRED07 from P14 contained 10 (71.4%) ST1424 and four (28.6%) ST80 isolates; sample VRED11 from P50 contained 13 (92.9%) ST1424 and one (7.1%) ST1659 isolate. A further rectal swab sample from P49 collected two days after VRED06 contained only ST1424, and a blood culture collected nine days later also contained ST1424. P9 had three rectal swab samples collected over 11 days and had positive blood cultures one month later, all samples contained ST80 only. Our finding of multiple strains in 27% of patients is in line with recent studies showing up to half of patients carry 2-4 different *E. faecium* strains, and within-patient diversity varies over time.^20,24,61,62^

**Table 2.** MLST Sequence Types Detected.

| Patient ID | Sample ID | Sample Type | Sample Date (days from start of study) | STs detected (n, %) | Maximum pairwise SNP distance within sample | Median (IQR) pairwise SNP distance within sample |
| --- | --- | --- | --- | --- | --- | --- |
| P2 | VRED01 | Rectal | 6 | 80 (14, 100) | 2 | 0 (0 - 1) |
| P6 | VRED16 | Rectal | 27 | 1424 (14, 100) | 2 | 0 (0 - 1) |
| P7 | VRED10 | Rectal | 18 | 80 (14, 100) | 2 | 0 (0 - 1) |
| P9 | VRED02 | Rectal | 6 | 80 (14, 100) | 2 | 0 (0 - 0) |
| P9 | VRED03 | Rectal | 8 | 80 (14, 100) | 0 | 0 (0 - 0) |
| P9 | VRED09 | Rectal | 17 | 80 (14, 100) | 0 | 0 (0 - 0) |
| P9 | VRED18 | Blood | 52 | 80 (1, 100) | NA | NA |
| P9 | VRED19 | Blood | 52 | 80 (1, 100) | NA | NA |
| P9 | VRED20 | Blood | 52 | 80 (1, 100) | NA | NA |
| P9 | VRED21 | Blood | 52 | 80 (1, 100) | NA | NA |
| P14 | VRED07 | Rectal | 17 | 1424 (10, 71.4)<br>80 (4, 26.6) | 2<br>2 | 1 (0-1)<br>2 (1-2) |
| P20 | VRED15 | Rectal | 27 | 80 (14, 100) | 3 | 0 (0 - 2) |
| P20 | VRED17 | Rectal | 30 | 80 (14, 100) | 3 | 1 (0 - 1) |
| P24 | VRED13 | Rectal | 26 | 1424 (14, 100) | 0 | 0 (0 - 0) |
| P33 | VRED12 | Rectal | 24 | 80 (14, 100) | 1 | 0 (0 - 0) |
| P34 | VRED04 | Rectal | 9 | 1424 (14, 100) | 0 | 0 (0 - 0) |
| P34 | VRED05 | Rectal | 14 | 1424 (14, 100) | 1 | 0 (0 - 0) |
| P49 | VRED06 | Rectal | 15 | 80 (10, 71.4)<br>1424 (3, 21.4)<br>789 (1, 7.1) | 2<br>1<br>NA | 0 (0 - 1)<br>0 (0 - 0)<br>NA |
| P49 | VRED08 | Rectal | 17 | 1424 (14, 100) | 2 | 0 (0 - 1) |
| P49 | VRED14 | Blood | 30 | 1424 (1, 100) | NA | NA |
| P50 | VRED11 | Rectal | 23 | 1424 (13, 92.9)<br>1659 (1, 7.1) | 3<br>NA | 1 (0 - 2)<br>NA |

### Genomic population structure of VREfm suggests recent transmission events

The chromosomes of the two strain-specific genome assemblies (Table S1) were used as references for short-read mapping within each sequence type. Within-patient diversity was low when genomes of the same sequence type were compared, generally differing by zero SNPs and a maximum pairwise difference of 3 SNPs (Table 2). Similarly, insertions, deletions, and plasmids were usually shared in genomes from the same patient. However, the presence of DEL3 (12 bp non-coding deletion) and DEL4 (11 bp deletion in a solute binding protein) were variable within 24 ST80 genomes from P20 with 0-2 differentiating SNPs (Figure 2). In genomes from P9 p1_VRED06-10 and p3_VRED06-10 were variably detected despite most genomes having no differentiating SNPs (Figure 2). Where multiple samples from the same patient were collected over time we found low (0-3 SNPs) accumulation of SNPs and no pattern in the prevalence of other genomic variants. Estimates of diversification rates in *E. faecium* from single colony sampling of national isolate collections suggest 7 mutations per year,^63^ other studies of longitudinal within-patient diversification have estimated higher rates of 12.6 – 128 mutations per year.^20,22,23^ The low SNP diversity identified in our one-month collection of carriage isolates is in keeping with these estimated mutation rates.

**Figure 2.**
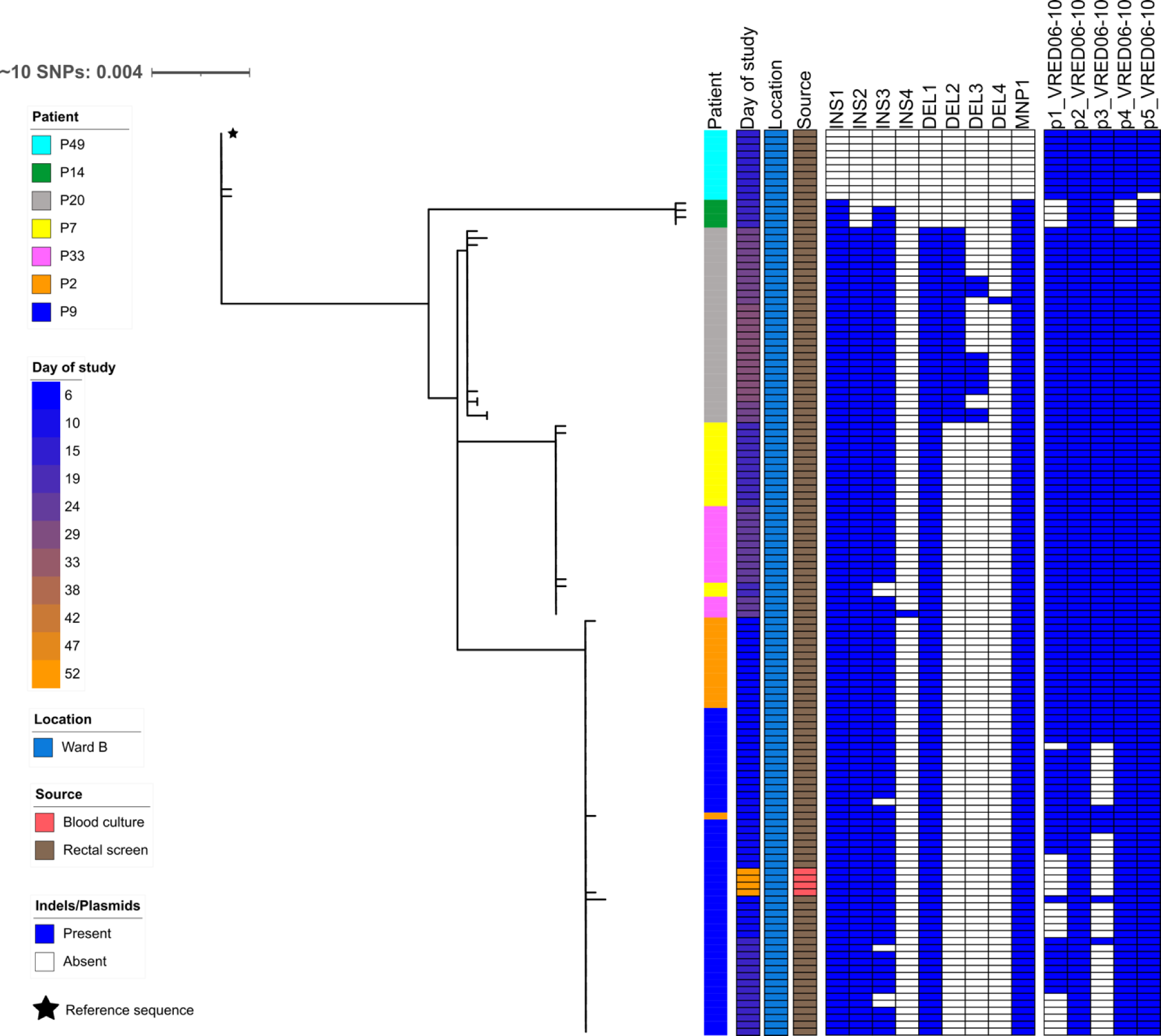
Phylogeny of ST80 isolates showing structured population with three patient specific clusters and two clusters indicating putative patient-patient transmission of VREfm. All ST80 isolates (n=130) mapped to VRED06-10 chromosome and phylogeny built on SNP sites (n=96) after removal of putative transposable and recombinant regions. Tree unrooted.

The ST80 genomes formed a well-structured population with five clear clusters each separated by >10 SNPs (Figure 2). Clustered genomes differed by 0-2 SNPs and were mostly from individual patients although two clusters included genomes from two different patients (patients P7 and P33, and P2 and P9). All the reference plasmids were detected in the P7 and P33 genomes and there were two insertions detected in three genomes. There was variation in detection of p1_VRED06-10 and p3_VRED06-10 plasmids within P9 genomes, although in P2 genomes all plasmids were detected.

Mapping of the ST1424 genomes showed a much more homogeneous population than in ST80 (Figure 3). Of the 97 ST1424 genomes, 69 had no SNPs and the remaining 28 had 1-2 SNPs differentiating them from the rest of the collection. The SNPs that were detected did not lead to any clear clustering of genomes, except for the 14 genomes from P6 which all carried a SNP in a penicillin-binding protein which differentiated them from the other ST1424 genomes. Two of the P6 genomes had further independent SNPs (one each) and another genome had lost p1_VRED06-02. No insertions were detected in the ST1424 collection, and of the six deletions found five were only in genomes from P49. p6_VRED06-02 was not detected in 14 P24, 14 P50, and two P49 genomes, while p1_VRED06-02 was note detected in five genomes from three patients.

**Figure 3.**
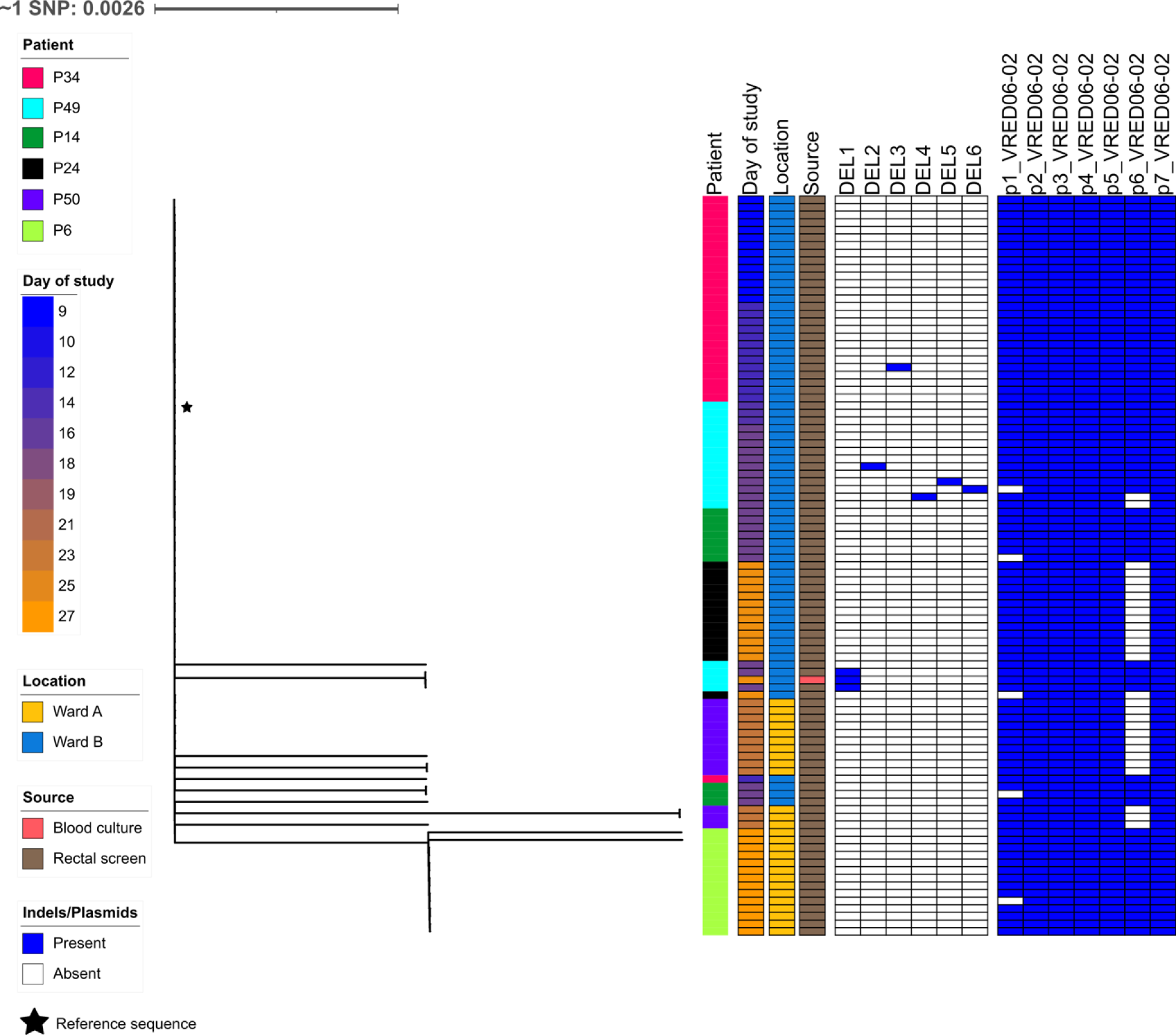
Unrooted phylogeny of ST1424 genomes showing homogeneous population suggestive of recent transmission outbreak. ST1424 genomes (n=97) mapped to VRED06-02 chromosome and phylogeny built on SNP sites (n=13) after removal of putative transposable and recombinant regions. Tree unrooted.

### Analysis of multiple VREfm colonies supports transmission resolution

A transmission network was constructed considering the phylogenetic placement of all 14 colony picks in each sample (Figure 4). The network supports transmission of ST80 between P2 and P9, and between P7 and P33, with P20 not linked to transmission. Epidemiological data supports transmission from P33 to P7 on Ward B, as P33 screened negative early in their admission and then screened positive six days after P7 (Figure 1, Figure 4). P9 and P2 screened positive on the same day - no shared rooms or bed spaces were identified as this was P2’s first day on Ward B so it is unclear where or when transmission may have occurred (Figure 1, Figure S1). P20 had two admissions during the study period, was negative at the end of first admission then screened positive on re-admission suggesting they may have become colonised outside of the hospital.

**Figure 4.**
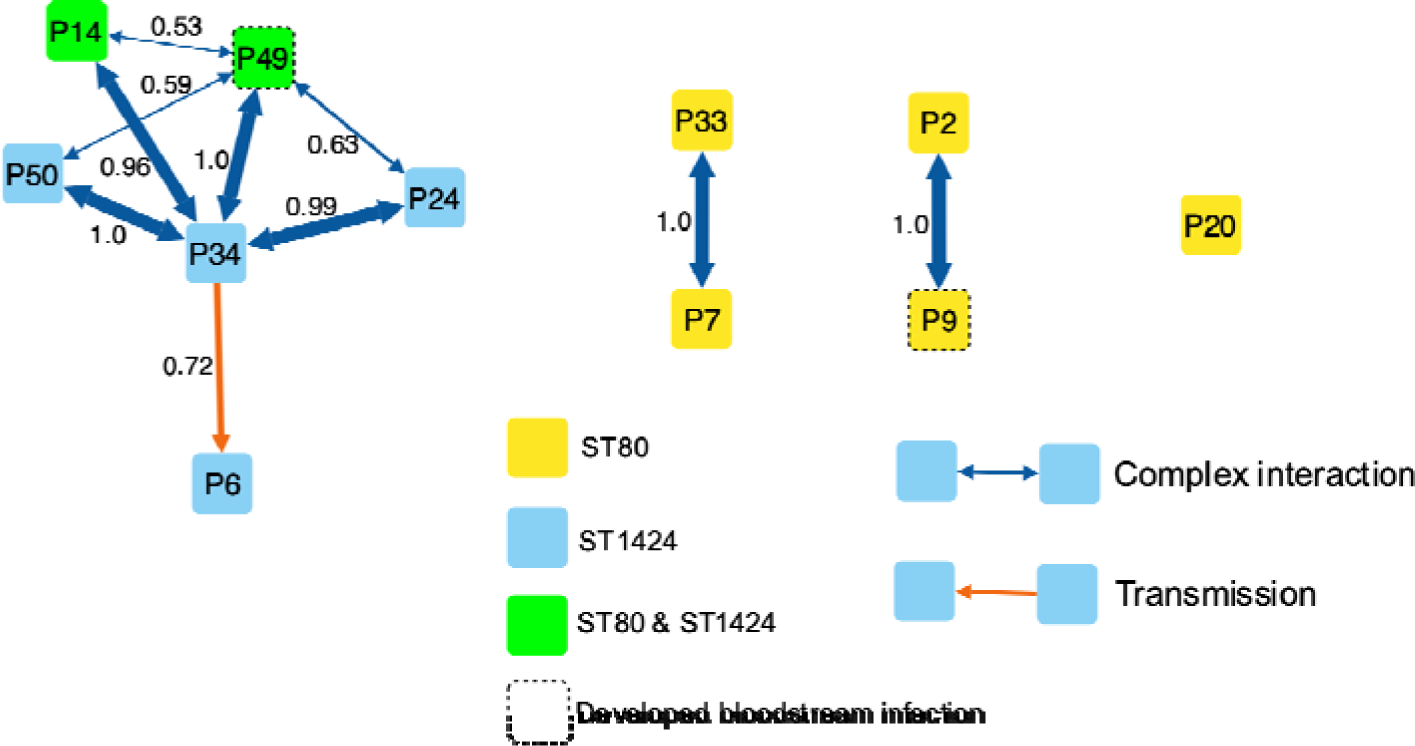
Phyloscanner transmission network. Each patient is represented by a node coloured by detection of the two outbreak STs. Edge thickness corresponds to fraction of Phyloscanner trees with given relationship, relationship fraction is printed alongside each edge, and edge colour based on type of relationship (orange, direct transmission; blue, transmission but direction unclear).

All ST1424 patients clustered together with P34 strongly linked to all patients and likely direct transmission to P6 (Figure 4). P34 was the first ST1424 identified on Ward B, P49 was positive six days later (having been negative earlier in admission), P14 was positive two days after that, and P24 was positive 9 days subsequently (Figure 1). P14 and P49 had ST1424 and ST80 in carriage samples, we did not identify sharing of the ST80 lineages in these patients suggesting there was no direct transmission between these two patients. On Ward A, P50 screened positive with ST1424 and ST1659 on day two of admission and P6 screened positive for ST1424 on day six. The ST1424 populations in P6 and P50 may derive from different hosts with P6 genomes all having a single SNP and P50 genomes having multiple different SNPs and lack the p6_VRED06-02 plasmid (Figures 3 and 4). P34 and P50 shared time on Ward A early in the study before either were known to be VREfm positive, but there is very limited overlap in time between P34 and P6 while both were in different wards (Figure 1). None of the patients with ST1424 shared a room or used a bed space previously used by an identified ST1424-positive carrier during their stay (Figure S1).

Analysing less than 14 colonies per sample produced fewer transmission links and lower confidence (Table S2, Figure S2). Sequencing more than 14 colonies would improve the detection of minor variants while increasing costs and turnaround time. There was low within-patient diversity in most patients in our study so this approach may not be required in every case. Alternatively, strain-resolved metagenomics directly on clinical samples or sweeps of selective culture growth may be more feasible.^64–66^ Further work is required to determine the optimum sampling strategy to support infection prevention and control investigations in healthcare settings.

### Plasmids were mostly ST-specific

VRED06-02 (ST1424 reference) contained seven plasmids, and VRED06-10 (ST80 reference) contained five plasmids. Plasmids in the two genomes were generally distinct, suggesting limited sharing between STs within P49 (Table S3).

We sought to identify carriage of similar plasmids in the entire collection by short read mapping (Table S4). Most plasmids were ST-specific with few examples of ST1424 genomes carrying plasmids from the ST80 reference, and *vice versa*. However, all ST80 genomes from P7 and P33 carried p7_VRED06-02 from ST1424, and almost all genomes appeared to carry p4_VRED06-10. We believe the hits against the ST1424 genomes are due to cross-mapping of reads from the related p4_VRED06-02 (Table S3). P7_VRED06-02 is unrelated to others in the collection (Table S3), but no close links to any ST1424-positive patients were identified for P7 and P33 (Figure 4).

### AMR gene load differs between closely related genomes

We next sought to determine the variability of AMR genes within the collection (Table 3 and Figure 5). In total 13 AMR genes were detected with three (*aac(6’)-Ii*, *msr(C)*, and *vanA*) present in all genomes, two (*aph(3’)-III* and *erm(B)*) in all but one genome, four (*ant(9)-Ia*, *dfrG*, *erm(A)*, and *tet(M)*) only in ST1424 or ST1659 genomes, two genes (*ant(6)-Ia* and *tet(S)*) found only in ST80 and ST789 genomes, and *tet(L)* found in a single ST1659 genome. The aminoglycoside resistance gene *aac(6’)-aph(2’’)* was variably present, found in 69.9% of all genomes.

**Figure 5.**
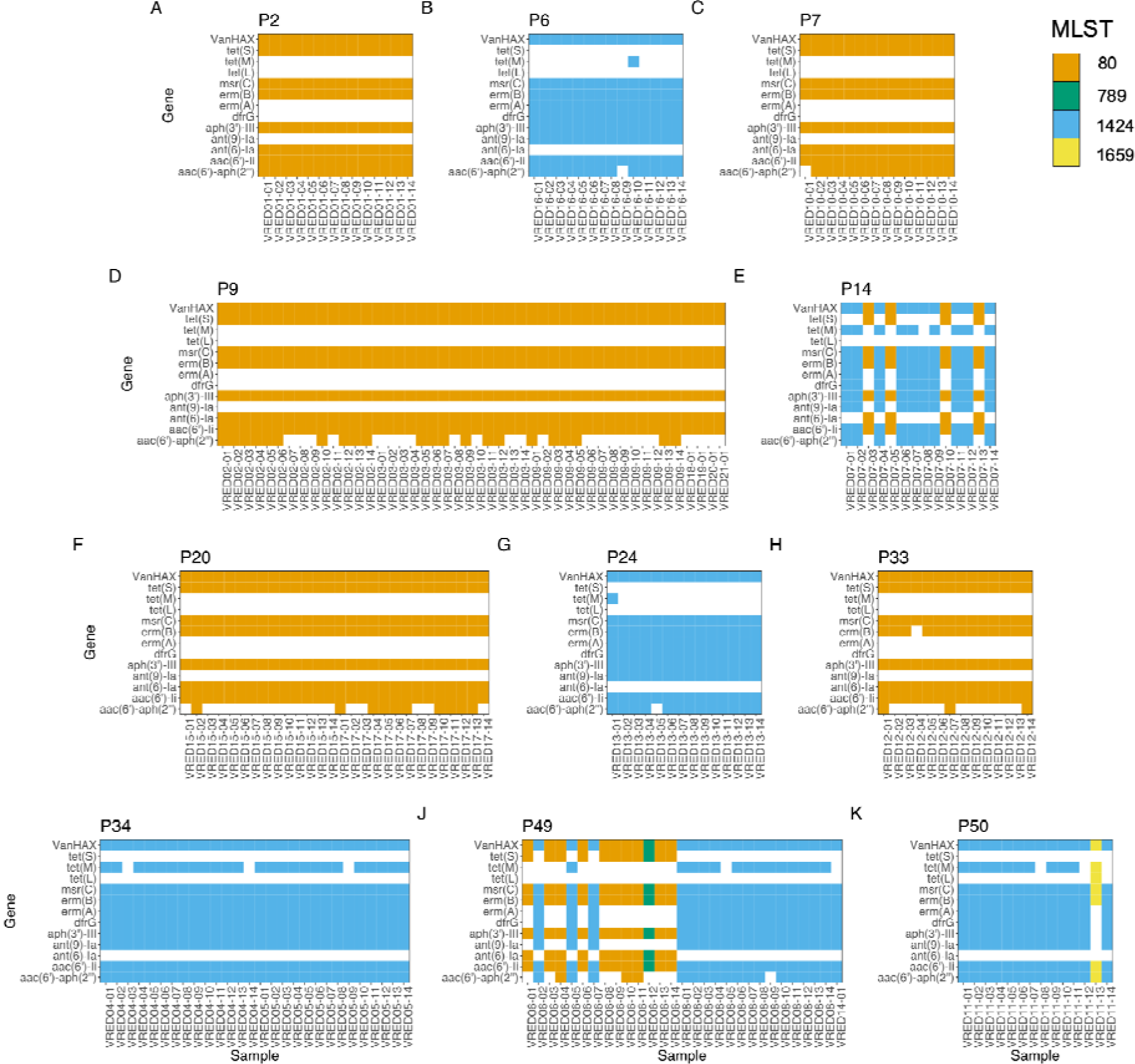
Detection of antimicrobial resistance genes. Panels represent different patients; resistance genes are plotted on the y-axis and isolates on the x-axis. Presence of a gene is represented by a filled square and coloured based on the MLST of the genome.

**Table 3.** Presence of AMR Genes.

| Gene | Phenotypic resistance | ST80, n = 130 |  | ST1424, n = 97 |  | All Genomes, n = 229 |  |
| --- | --- | --- | --- | --- | --- | --- | --- |
|  |  | n (%) | Genetic element | n (%) | Genetic element | n (%) | Summary |
| *aac(6')-aph(2'')* | Amikacin, Gentamicin, Kanamycin, Streptomycin, Tobramycin | 65 (50.0) | p1_VRED06-10 | 94 (96.9) | p1_VRED06-02 | 160 (69.9) | Variable in ST80/ST1424/ST789 |
| *aac(6')-li* | Gentamicin, Tobramycin | 130 (100) | Chromosome | 97 (100) | Chromosome | 229 (100) | All genomes |
| *ant(6)-la* | Streptomycin | 130 (100) | p3_VRED06-10 | 0 (0) | NA | 131 (57.2) | All ST80/ST789 |
| *ant(9)-la* | Spectinomycin | 0 (0) | NA | 97 (100) | Chromosome | 97 (42.4) | All ST1424 |
| *aph(3')-III* | Amikacin, Kanamycin, Neomycin | 130 (100) | p3_VRED06-10 | 97 (100) | p2_VRED06-02 | 228 (99.6) | All ST80/ST789/ST1424 |
| *dfrG* | Trimethoprim | 0 (0) | NA | 97 (100) | Chromosome | 97 (42.4) | All ST1424 |
| *erm(A)* | Clindamycin, Erythromycin, Quinupristin | 0 (0) | NA | 97 (100) | Chromosome | 97 (42.4) | All ST1424 |
| *erm(B)* | Clindamycin, Erythromycin, Quinupristin | 129 (99.2) | p1_VRED06-10, p3_VRED06-10 | 97 (100) | p2_VRED06-02 | 228 (99.6) | All except one ST80 genome |
| *msr(C)* | Erythromycin, Quinupristin | 130 (100) | Chromosome | 97 (100) | Chromosome | 229 (100) | All genomes |
| *tet(L)* | Doxycycline, Tetracycline | 0 (0) | NA | 0 (0) | NA | 1 (0.4) | Only ST1659 |
| *tet(M)* | Doxycycline, Minocycline, Tetracycline | 0 (0) | NA | 60 (61.9) | Chromosome | 61 (26.6) | Variable in ST1424/ST1659 |
| *tet(S)* | Doxycycline, Minocycline, Tetracycline | 130 (100) | p3_VRED06-10 | 0 (0) | NA | 131 (57.2) | All ST80/ST789 |
| *VanA* | Teicoplanin, Vancomycin | 97 (100) | p2_VRED06-02 | 97 (100) | p2_VRED06-02 | 229 (100) | All genomes |

Tetracycline resistance gene *tet(M)* was identified on the chromosome of VRED06-02 as part of Tn*6944* (Figure S3A). *tet(M)* was identified in 62.2% of ST1424 and ST1659 genomes, excision of Tn*6944* may be responsible for this variable presence. We identified variable within-patient presence of *tet(M)* and no other tetracycline resistance genes (Figure 5), phenotypic susceptibility pattern would therefore differ based on which colony was picked. However, tetracyclines are not generally used for treatment of enterococcal human infections so the clinical impact may be limited. Similar variable presence of the vancomycin resistance element within patients has been described elsewhere and could lead to inappropriate use of vancomycin when the patient harbours a resistant subpopulation.^20,21,67,68^ Our study only included vancomycin resistant isolates, so cannot resolve the potential role of variable vancomycin resistance carriage within patients or in transmission networks. Gain and loss of vancomycin resistance has been described in regional networks over periods of years^63^.

*aac(6’)-aph(2’’)* was present on p1_VRED06-02 (ST80) and p1_VRED06-10 (ST1424). *aac(6’)-aph(2’’)* was not detected in any ST80 genomes that were p1_VRED06-02 negative, although only 39.8% (n=43) of genomes that carried this plasmid also carried *aac(6’)-aph(2’’)*. In p1_VRED06-02, two copies of *aac(6’)-aph(2’’)* were surrounded by insertion sequences IS*256*, IS*1216*, and IS*3,* providing multiple mechanisms of excision. In ST1424 *aac(6’)-aph(2’’)* was detected in 97.8% (n=90) genomes with p1_VRED06-10. In p1_VRED06-10, *aac(6’)-aph(2’’)* was surrounded by two copies of IS*256* similarly to Tn*6218*, although the transposition machinery was missing (Figure S3B).^69^ Another four ST1424 genomes carried *aac(6’)-aph(2’’)* but not p1_VRED06-10 (Table 3 and Figure 5). Short read assemblies could not resolve the environment of *aac(6’)-aph(2’’)*, but in three cases *aac(6’)-aph(2’’)* co-located with an IS*3* gene suggesting mobilisation to another transposable element. The impact on phenotype is unclear – all genomes carried *aac(6’)-Ii* and *aph(3’)-III* which together confer high-level resistance to the clinically relevant aminoglycosides amikacin and gentamicin, so the loss of *aac(6’)-aph(2’’)* may be more efficient for the cell without an overt change in antibiotic susceptibility. Both Tn*6994* and Tn*6218* were first characterised in *C. difficile*, highlighting transmission of AMR elements between nosocomial pathogens as recently described.^70^

The tetracycline resistance gene *tet(L)* was identified in a single ST1659 genome, the gene was co-located with *tet(M)* on a 30 kb contig that was similar to Tn*6248* from *E. faecium* over ∼19 kb (Figure S3C).

We recognise some limitations. Around 60% of *E. faecium* carriers can be linked to nosocomial transmission from other patients or reservoirs in the hospital environment.^61,71–74^ Our study did not include environmental samples, and although patients were mostly located in individual rooms bathroom facilities were shared posing a significant environmental reservoir for VREfm. Also, we relied on direct plating to solid VREfm screening agar for inclusion in our study.

Previous studies have shown a sensitivity of 58-96% for this approach, rising to 97-100% with a pre-enrichment step.^75–77^

A proactive sequence-based surveillance approach should avoid large infection outbreaks, and reduce ward closure costs and the clinical impact of invasive disease.^78–81^ In our setting, an outbreak of VREfm was suspected 3 weeks after the study collection period when P9 and P49 developed bloodstream infection concurrently but this was many weeks after VREfm transmission had likely occurred (Figure 1 and Figure 4). Due to our study’s retrospective nature, we could not use the findings from sequencing to directly influence patient care.

To conclude, by taking account of within-patient diversity in VREfm carriage populations we identified transmission links between patients that could supplement efforts to control transmission within hospitals. We also show that diversity exists not just at the level of SNPs – AMR gene presence/absence, indels, and plasmid presence all vary within and between patients. Accounting for within-patient diversity is important for resolving VREfm transmission using WGS-based investigations.

## Data Availability

Sequence data have been uploaded to the NCBI under BioProject PRJNA877253

https://www.ncbi.nlm.nih.gov/bioproject/PRJNA877253

## ACKNOWLEDGEMENTS

The authors would like to thank all staff at the Department for Medical Microbiology, Royal Infirmary of Edinburgh for supporting this study. The authors acknowledge the Research/Scientific Computing teams at The James Hutton Institute and NIAB for providing computational resources and technical support for the “UK’s Crop Diversity Bioinformatics HPC” (BBSRC grant BB/S019669/1), use of which has contributed to the results reported within this paper. Bioinformatics and Computational Biology analyses were further supported by the University of St Andrews Bioinformatics Unit which is funded by a Wellcome Trust ISSF award [grant 105621/Z/14/Z].

## TRANSPARENCY DELCARATIONS

The authors declare no competing interests.

