## Supplementary file for "Consideration of within-patient diversity highlights transmission pathways and antimicrobial resistance gene variability in vancomycin resistant *Enterococcus faecium*"

**Contents**

Table S2 Transmission network scores for different colony thresholds – pg 1

Table S3 Mash distance of reference isolate plasmids – pg 2

Table S4 Plasmid detection in genome collection – pg 3

Figure S1 Map of patient locations at time of screening positive for VREfm – pg 5

Figure S2 Effect of different sampling strategies on transmission inference – pg 6

Figure S3 Comparison of AMR elements to previously described transposons – pg 8

**Table S2 –** Transmission network scores for different colony thresholds

|  | **3 Colonies** | **5 Colonies** | **10 Colonies** | **14 Colonies** |
| --- | --- | --- | --- | --- |
| **Total Colonies** | 30 | 50 | 100 | 229 |
| **Transmissions detected** | 9 | 9 | 10 | 11 |
| **Transmission confidence, median (min-max)** | 0.84  (0.58-0.99.0) | 0.73  (0.57-1.0) | 0.90  (0.53-1.0) | 0.98  (0.53-1.0) |

**Table S3** – Mash distance of reference isolate plasmids

|  | **p1_VRED06-02** | **p2_VRED06-02** | **p3_VRED06-02** | **p4_VRED06-02** | **p5_VRED06-02** | **p6_VRED06-02** | **p7_VRED06-02** | **p1_VRED06-10** | **p2_VRED06-10** | **p3_VRED06-10** | **p4_VRED06-10** | **p5_VRED06-10** |
| --- | --- | --- | --- | --- | --- | --- | --- | --- | --- | --- | --- | --- |
| **p1_VRED06-02** | 0.00 | 0.15 | 0.13 | 1.00 | 1.00 | 1.00 | 1.00 | 0.06 | 0.20 | 0.11 | 1.00 | 1.00 |
| **p2_VRED06-02** | 0.15 | 0.00 | 0.17 | 0.30 | 0.16 | 0.30 | 0.16 | 0.16 | 0.09 | 0.09 | 0.23 | 0.30 |
| **p3_VRED06-02** | 0.13 | 0.17 | 0.00 | 1.00 | 1.00 | 1.00 | 1.00 | 0.11 | 0.18 | 0.09 | 1.00 | 1.00 |
| **p4_VRED06-02** | 1.00 | 0.30 | 1.00 | 0.00 | 0.18 | 1.00 | 1.00 | 1.00 | 1.00 | 0.24 | 0.05 | 0.16 |
| **p5_VRED06-02** | 1.00 | 0.16 | 1.00 | 0.18 | 0.00 | 0.15 | 1.00 | 1.00 | 1.00 | 0.24 | 0.15 | 0.11 |
| **p6_VRED06-02** | 1.00 | 0.30 | 1.00 | 1.00 | 0.15 | 0.00 | 0.17 | 1.00 | 1.00 | 0.26 | 1.00 | 1.00 |
| **p7_VRED06-02** | 1.00 | 0.16 | 1.00 | 1.00 | 1.00 | 0.17 | 0.00 | 1.00 | 1.00 | 0.24 | 1.00 | 1.00 |
| **p1_VRED06-10** | 0.06 | 0.16 | 0.11 | 1.00 | 1.00 | 1.00 | 1.00 | 0.00 | 1.00 | 0.17 | 1.00 | 1.00 |
| **p2_VRED06-10** | 0.20 | 0.09 | 0.18 | 1.00 | 1.00 | 1.00 | 1.00 | 1.00 | 0.00 | 1.00 | 1.00 | 1.00 |
| **p3_VRED06-10** | 0.11 | 0.09 | 0.09 | 0.24 | 0.24 | 0.26 | 0.24 | 0.17 | 1.00 | 0.00 | 0.23 | 0.26 |
| **p4_VRED06-10** | 1.00 | 0.23 | 1.00 | 0.05 | 0.15 | 1.00 | 1.00 | 1.00 | 1.00 | 0.23 | 0.00 | 0.18 |
| **p5_VRED06-10** | 1.00 | 0.30 | 1.00 | 0.16 | 0.11 | 1.00 | 1.00 | 1.00 | 1.00 | 0.26 | 0.18 | 0.00 |

Coloured based on similarity: ≤0.01, green; ≤0.05, yellow; ≤0.1, blue.

**Table S4** – Plasmid detection in genome collection

| Patient | STs (n) | ST1424 Reference Plasmids | | | | | | | ST80 Reference Plasmids | | | | |
| --- | --- | --- | --- | --- | --- | --- | --- | --- | --- | --- | --- | --- | --- |
|  |  | p1_VRED06-02 | p2_VRED06-02 | p3_VRED06-02 | p4_VRED06-02 | p5_VRED06-02 | p6_VRED06-02 | p7_VRED06-02 | p1_VRED06-10 | p2_VRED06-10 | p3_VRED06-10 | p4_VRED06-10*^a^* | p5_VRED06-10 |
| P34 | 1424 (28) | 28 (100) | 28 (100) | 28 (100) | 28 (100) | 28 (100) | 28 (100) | 28 (100) | 0 (0) | 0 (0) | 0 (0) | 28 (100) | 0 (0) |
| P6 | 1424 (14) | 13 (92.9) | 14 (100) | 14 (100) | 14 (100) | 14 (100) | 14 (100) | 14 (100) | 0 (0) | 0 (0) | 0 (0) | 14 (100) | 0 (0) |
| P24 | 1424 (14) | 13 (92.9) | 14 (100) | 14 (100) | 14 (100) | 14 (100) | 0 (0) | 14 (100) | 0 (0) | 0 (0) | 0 (0) | 14 (100) | 0 (0) |
| P50 | 1424 (13) | 13 (100) | 13 (100) | 13 (100) | 13 (100) | 13 (100) | 0 (0) | 13 (100) | 0 (0) | 0 (0) | 0 (0) | 13 (100) | 0 (0) |
|  | 1659 (1) | 1 (100) | 1 (100) | 0 (0) | 1 (100) | 1 (100) | 0 (0) | 1 (100) | 0 (0) | 0 (0) | 0 (0) | 1 (100) | 0 (0) |
| P14 | 1424 (10) | 8 (80.0) | 10 (100) | 10 (100) | 10 (100) | 10 (100) | 10 (100) | 10 (100) | 0 (0) | 0 (0) | 0 (0) | 10 (100) | 0 (0) |
|  | 80 (4) | 0 (0) | 0 (0) | 0 (0) | 0 (0) | 0 (0) | 0 (0) | 0 (0) | 0 (0) | 4 (100) | 4 (100) | 0 (0) | 4 (100) |
| P49 | 1424 (18) | 17 (94.4) | 18 (100) | 18 (100) | 18 (100) | 18 (100) | 16 (88.9) | 18 (90.9) | 0 (0) | 0 (0) | 0 (0) | 18 (100) | 0 (0) |
|  | 80 (10) | 0 (0) | 0 (0) | 0 (0) | 0 (0) | 0 (0) | 0 (0) | 0 (0) | 10 (100) | 10 (100) | 10 (100) | 10 (100) | 9 (90.9) |
|  | 789 (1) | 0 (0) | 0 (0) | 0 (0) | 0 (0) | 0 (0) | 0 (0) | 0 (0) | 1 (100) | 1 (100) | 1 (100) | 1 (100) | 1 (100) |
| P7 | 80 (14) | 0 (0) | 0 (0) | 0 (0) | 0 (0) | 0 (0) | 0 (0) | 14 (100) | 14 (100) | 14 (100) | 14 (100) | 14 (100) | 14 (100) |
| P20 | 80 (28) | 0 (0) | 0 (0) | 0 (0) | 0 (0) | 0 (0) | 0 (0) | 0 (0) | 28 (100) | 28 (100) | 28 (100) | 28 (100) | 28 (100) |
| P2 | 80 (14) | 0 (0) | 0 (0) | 0 (0) | 0 (0) | 0 (0) | 0 (0) | 0 (0) | 14 (100) | 14 (100) | 14 (100) | 14 (100) | 14 (100) |
| P33 | 80 (14) | 0 (0) | 0 (0) | 0 (0) | 0 (0) | 0 (0) | 0 (0) | 14 (100) | 14 (100) | 14 (100) | 14 (100) | 14 (100) | 14 (100) |
| P9 | 80 (46) | 0 (0) | 0 (0) | 0 (0) | 0 (0) | 0 (0) | 0 (0) | 0 (0) | 28 (60.9) | 46 (100) | 10 (21.7) | 46 (100) | 46 (100) |

*^a^* p4_VRED06-10 is shorter than but homologous to p4_VRED06-02, the matches in P6, P24, and P34, and ST1424/1659 P14, P49, and P50 genomes are likely false positives

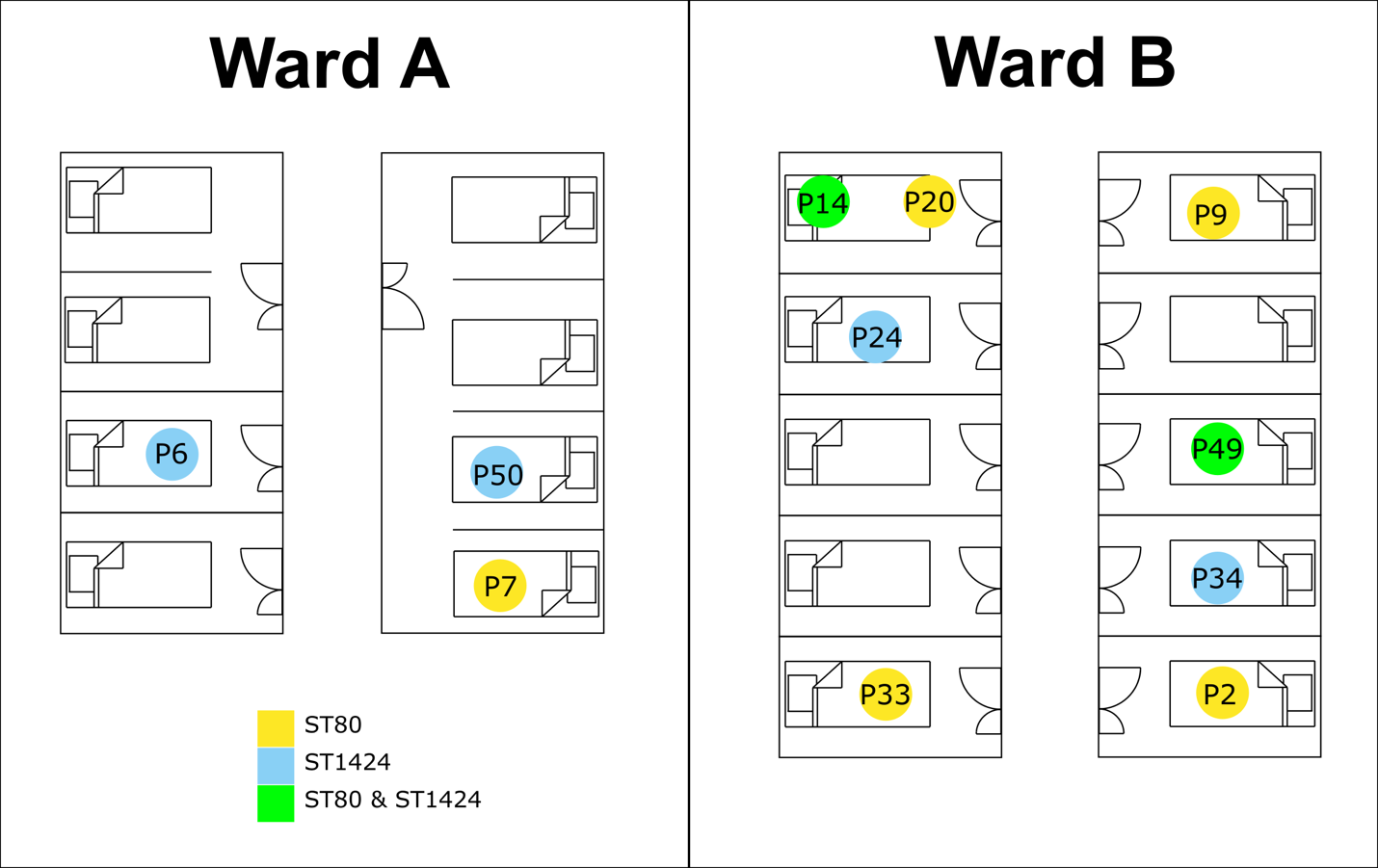

**Figure S1** – Map of patient locations at time of screening positive for VREfm

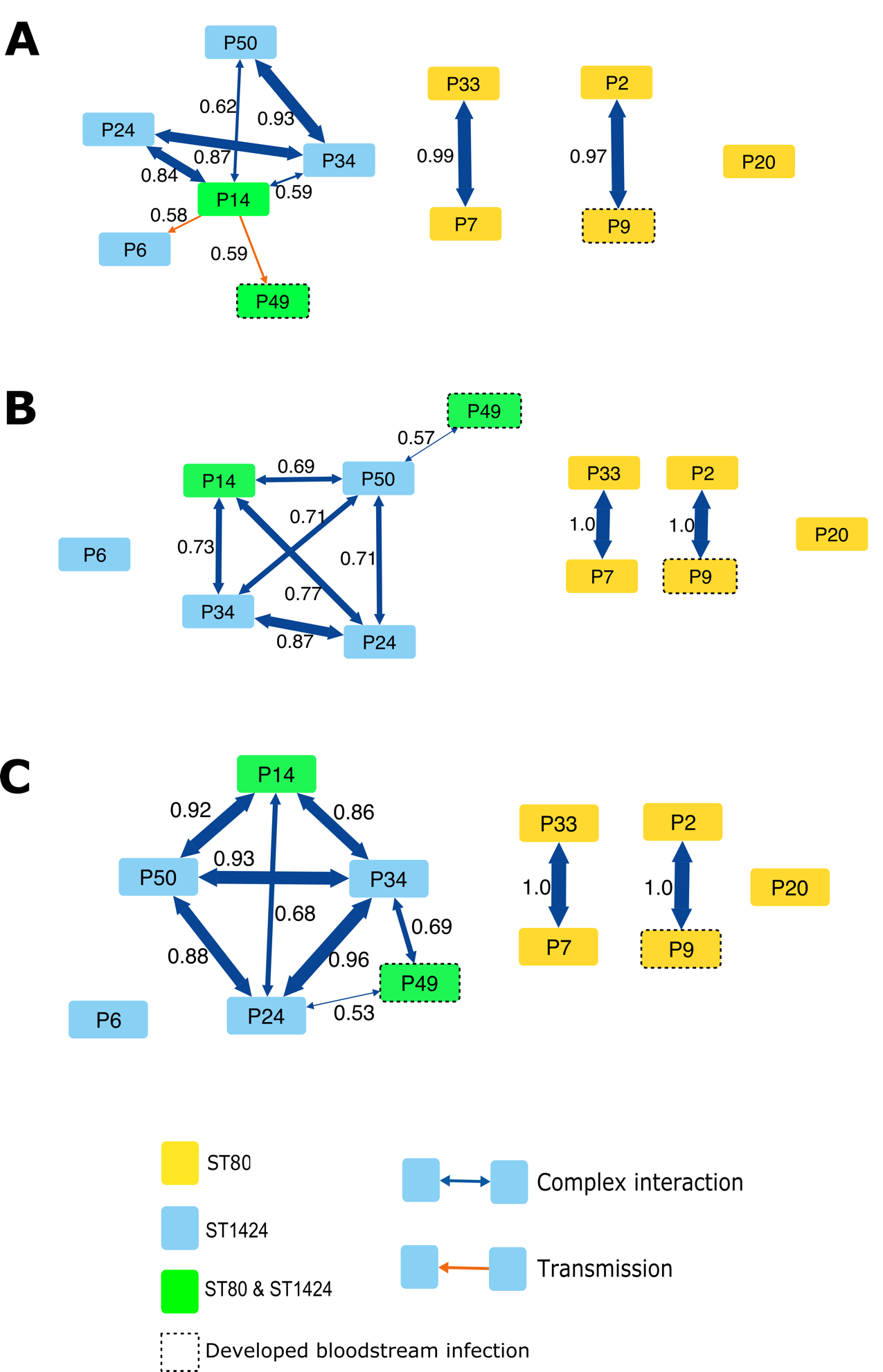

**Figure S2** – Effect of different sampling strategies on transmission inference. Phyloscanner transmission networks for 3 (A), 5 (B), and 10 (C) colony picks. Edge thickness corresponds to fraction of Phyloscanner trees with given relationship, relationship fraction is printed alongside each edge, and edge colour based on type of relationship (orange, direct transmission; blue, transmission but direction unclear).

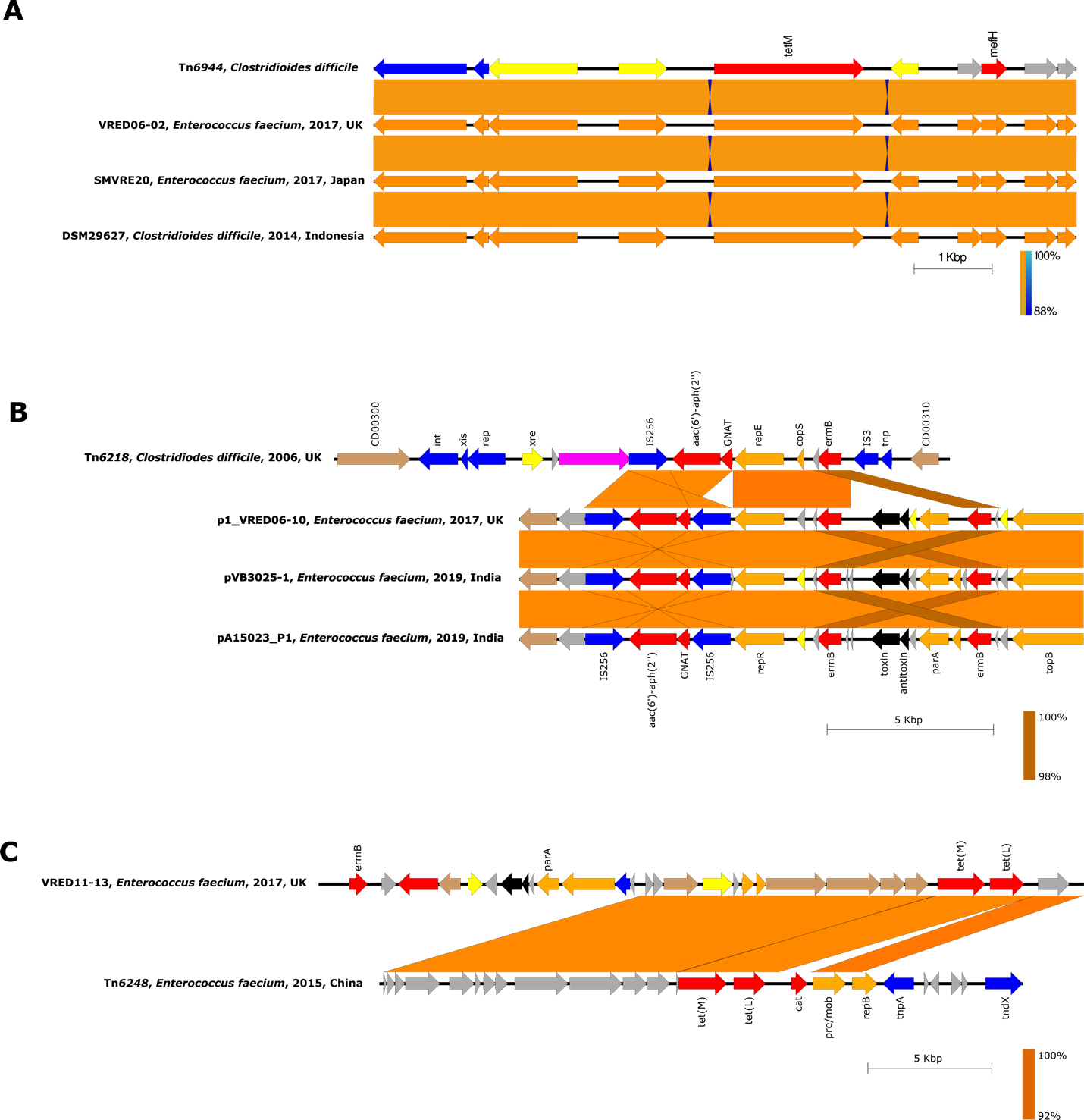

**Figure S3** – Comparison of AMR elements to previously described transposons. Genome comparison of (A) *tet(M)*, (B) *aac(6’)-aph(2’’)*, and (C) *tet(L)* elements to published examples. Transposon or strain identification, species, year, and country of first identification are given where available. Coding sequences are coloured based on inferred function: AMR, red; transposon, blue; replication, orange; regulation, yellow; toxin/antitoxins, black; hypothetical, grey; pink, surface-associated; other, brown.
